# Artificial Intelligence Model for Analyzing Colonic Endoscopy Images to Detect Changes Associated with Irritable Bowel Syndrome

**DOI:** 10.1101/2022.05.09.22274876

**Authors:** Kazuhisa Tabata, Hiroshi Mihara, Sohachi Nanjo, Iori Motoo, Takayuki Ando, Akira Teramoto, Haruka Fujinami, Ichiro Yasuda

## Abstract

**Background/Aims:** IBS is not an organic disease, and the patients typically show no abnormalities in lower gastrointestinal endoscopy. Recently, biofilm formation has been visualized by endoscopy, and the ability of endoscopy to detect microscopic changes due to dysbiosis and microinflammation has been reported. In this study, we investigated whether an Artificial Intelligence (AI) colon image model can detect IBS without biofilsm.

**Methods:** Study subjects were identified based on electronic medical records and categorized as IBS (group I; n=11), IBS with predominant constipation (IBS-C; group C; n=12), and IBS with predominant diarrhea (IBS-D; group D; n=12). Colonoscopy images from IBS patients and from asymptomatic healthy subjects (group N; n=88) were obtained. Google Cloud Platform AutoML Vision (single-label classification) was used to construct AI image models to calculate sensitivity, specificity, predictive value, and AUC. A total of 2479, 382, 538, and 484 images were randomly selected for groups N, IBS, IBC-C and IBS-D groups, respectively.

**Results:** The AUC of the model discriminating between group N and group I was 0.95. Sensitivity, specificity, positive predictive value, and negative predictive value of group I detection were 30.8%, 97.6%, 66.7%, and 90.2%, respectively. The overall AUC of the model discriminating between groups N, C, and D was 0.83; sensitivity, specificity, and positive predictive value of group N were 87.5%, 46.2%, and 79.9%, respectively.

**Conclusions:** Using the image AI model, colonoscopy images of IBS could be discriminated from healthy subjects at AUC 0.95. Prospective studies are needed to further validate whether this externally validated model has similar diagnostic capabilities at other facilities and whether it can be used to determine treatment efficacy.

## Introduction

Irritable bowel syndrome (IBS) affects about 10% of the Western population, and its prevalence is increasing annually[1, 2]. Patients with IBS frequently experience abdominal pain and changes in stool habits, but often exhibit no abnormalities in immediate diagnostic tests or lower gastrointestinal endoscopy[3]. Recent evidence indicates that aspects of Western lifestyles, such as frequent antibiotic therapy that alter the microbiota may be involved in development of IBS. Biofilm formation is a unique mode of microbial growth [4] and polymicrobial biofilms have been implicated in several gastrointestinal disorders[5, 6]. In a recent study, biofilm formation by *E. coli* and *Ruminococcus gnavus* in the terminal ileum to the ascending colon was seen in 60% of IBS cases, and close observation revealed changes in areas where biofilms formed[7]. Imaging artificial intelligence models (AI) have been developed to detect lower gastrointestinal tract lesions in real time, and several models have already been clinically applied[3-5]. Complex AI models such as those developed for imaging applications require deep learning algorithms and generally can only be built using Python libraries and require programming expertise. Although relatively few physicians have such skills, tools such as Google Cloud Platform (Google Inc. Mountain View, CA Available at: http://cloud.google.com/vision/. Accessed 13 Feb 2022) now allow the building of AI models without such programming expertise, and thus the application of AI models in the medical field is likely to expand[6]. In fact, use of AI in the pathological diagnosis of infertile spermatozoa and otolaryngology imaging has already been reported[7, 8].

Typically, training datasets are needed to develop AI, but such datasets are not available for functional gastrointestinal diseases, which do not show abnormalities even on endoscopy. However, inclusion of additional information such as the presence or absence of symptoms in training datasets used to develop AI models may allow detection of minute changes in the colon that cannot be detected by human observers. The purpose of this study was to determine whether image AI models can differentiate between different types of IBS and healthy colonoscopic images in real-world clinical practice using Google cloud AutoML Vision.

## Materials and Methods

The study protocol was approved by the Ethics Committee of the University of Toyama Hospital (Approval No. R2021032). All methods were conducted in accordance with relevant guidelines and regulations and the Declaration of Helsinki. The research plan was presented to the Ethics Committee with the condition that the opt-out policy, which allows patients and their relatives to refuse to participate in the study, is clearly stated on the University of Toyama Hospital website. For use in real-world IBS patients, patients were identified not by the ROME criteria, but based on disease names recorded for insurance purposes between January 2010 and December 2020. These names included “Irritable bowel syndrome (group I), “constipated irritable bowel syndrome (group C),” and “diarrhea irritable bowel syndrome (group D)”. Colonoscopy images for these patients and images of patients having abnormal health examination but no evidence of abnormalities on endoscopy (Olympus CF-HQ290Z and PCF-H290Z) (group N) were obtained from the endoscopy reporting system. Normal light images of the terminal ileum, rectal inversion and anus, narrow band or dye-spread images were excluded. A total of 20 to 40 images were used, with about 5 images for each region per patient. The N group had 88 patients and 2,479 images were used. Group I had 11 patients and 382 images were used. Group C had 12 patients and 538 images were used. Group D had 12 patients and 484 images were used. While the larger the number of patients, the more accurate the model can be, a minimum of 100 images for each label is sufficient to construct the model, and since a certain degree of accuracy was obtained with the number of patients and number of images are considered sufficient to construct this model.

In this study we used annotation and Algorithm Generation using Google Cloud AutoML Vision from the Google Cloud Platform (GCP) (Google, Inc.). Four labels were defined as Group N, I, C and D in the training dataset (single label classification). Two models were produced that differentiated IBS from healthy or differentiated IBS-C, IBS-D, and healthy. This process was done entirely by a single physician (HM).

### Artificial Neural Network Programming, Training and External Validation

The Google Cloud AutoML Vision platform was used to automatically and randomly select training set images (80%), validation set images (10%), and test set images (10%) from the dataset for algorithm training processes. Since these images are independent of each other, even external validation can be performed. A total of 16 nodes (2 hours) were used to train the algorithm. AutoML Vision does provide metrics: positive predictive values and sensitivity to stated thresholds, and area under the curve (AUC).

For each model, we also generated a confusion matrix that cross-references the true labels against the labels predicted by the deep learning mode[9]. Using the extracted binary diagnostic accuracy data, we created a contingency table showing the calculated values for specificity at a threshold of 0.5. The contingency table showed results for true positive, false positive, true negative, and false negative.

### Data Sharing

The image data used in Google Cloud AutoML Vision is owned by HM and can be used in other studies upon receiving approval from the Ethics Committee.

## Results

### Healthy (Group N) vs. IBS (Group I)

A comparison of patients in Group N (Healthy) for whom endoscopy showed no apparent abnormalities and Group I (patients with IBS) showed that the average precision (positive predictive value), precision and recall of the algorithm was 94.6%, 88.78% and 88.78%, respectively, based on automated training and testing using the AI model developed (Figure 1). Precision recall curves were generated for each individual label as well as for the algorithm overall. We adopted a threshold value of 0.5 to yield balanced precision and recall. The AUC of the model to discriminate group N and group I and the confusion matrix are shown in Table 1. The total AUC was 0.95 (group I AUC 0.48, group N AUC 0.97) and the sensitivity, specificity, positive predictive value and negative predictive value of group I detection were 30.8%, 97.6%, 66.7%, and 90.23%, respectively. We found that the confusing rate for group I and group N was 69% and 2%, respectively. Representative images from endoscopy for patients with high IBS scores (Figure 2) and high normal scores are shown (Figure 3).

**Fig. 1.**
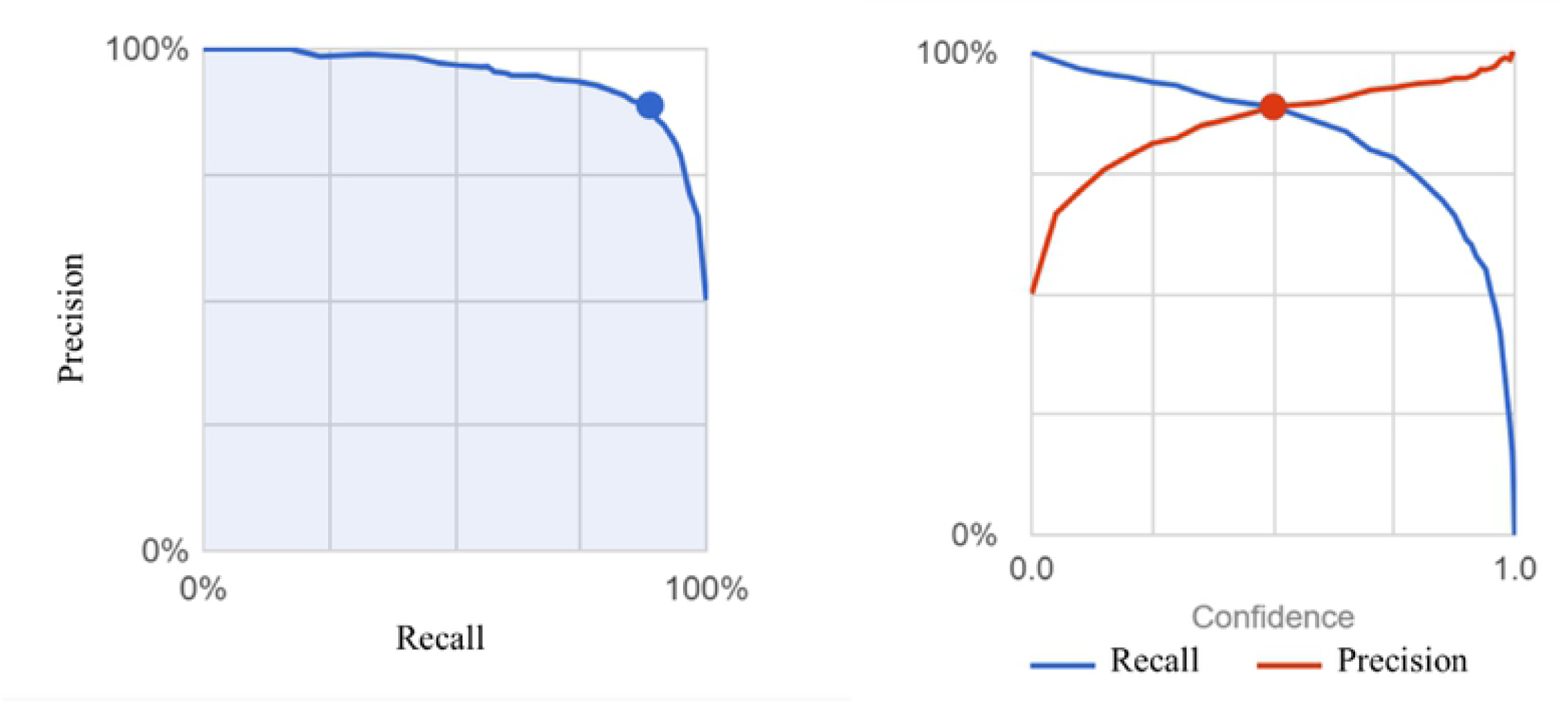
Positive predictive values, recall, and area under the curve (AUC) for a threshold value of 0.5 in IBS colon.

**Fig. 2.**
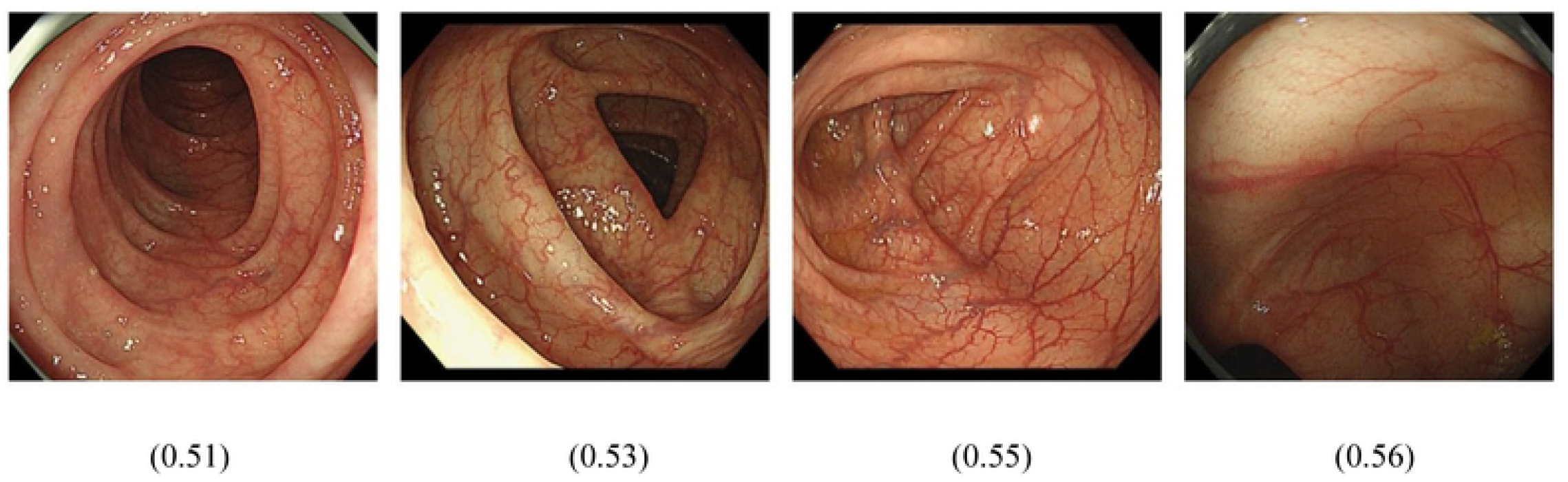
Images that scored relatively high (0 to 1) in the colon image AI model for detecting IBS disease names that were recorded for insurance purposes are shown.

**Fig. 3.**
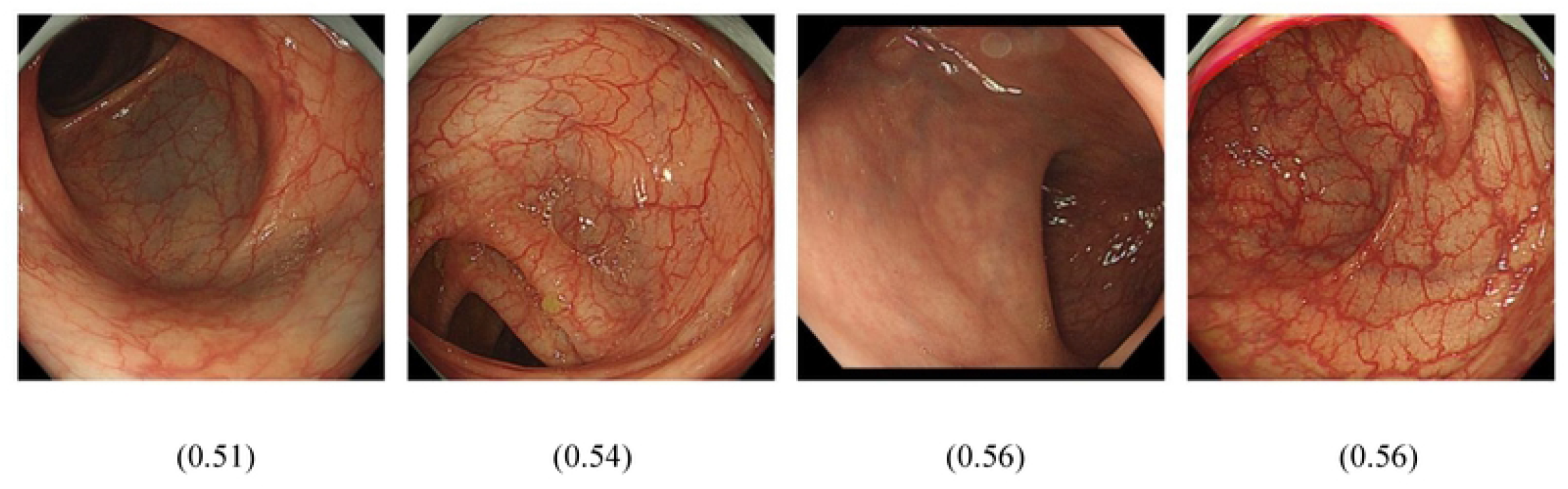
Images that scored relatively high (0 to 1) in the colon image AI model for detecting healthy individuals are shown.

**Table 1.**
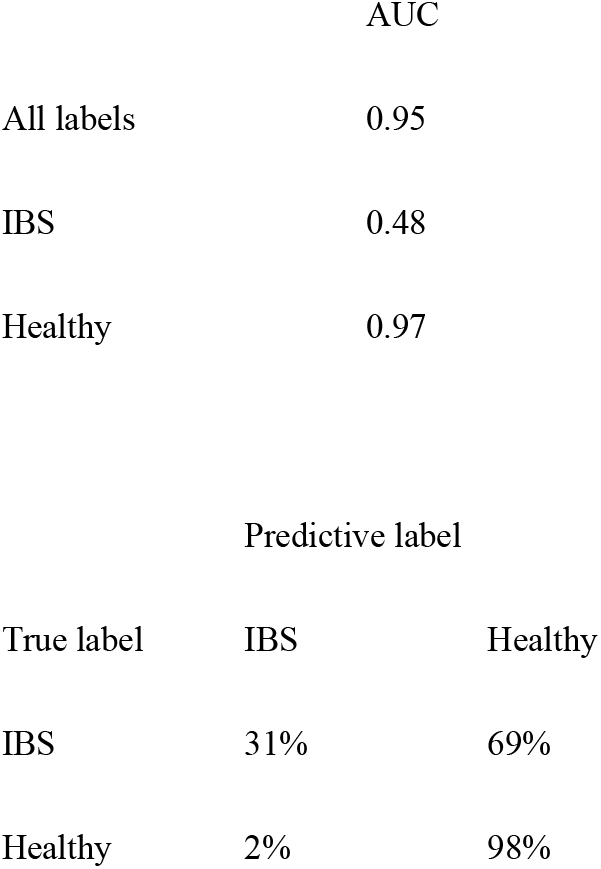
IBS model of the colon; AUC and contingency tables

### Healthy (Group N) vs. IBS-C (Group C) and IBS-D (Group D)

Next, images from Group N were compared with images from endoscopy for patients in Group C (IBS with constipation) and Group D (IBS with diarrhea). For these groups, the average precision (positive predictive value), precision, and recall of the algorithm were 83.2%, 77.71%, and 67.97%, respectively, based on automated training and testing (Figure 4). The precision recall curves and the threshold value were set as described above. The AUC of the model to discriminate among the groups and the confusion matrix are shown in Table 2. The total AUC was 0.83 (0.90 for group N, 0.45 for group C and 0.60 for group D) and the sensitivity, specificity, and positive predictive value of group N were 87.5%, 46.2%, and 79.9%, respectively. The confusing rate for group N, group D and group C was 12%, 51% and 66%, respectively.

**Fig. 4.**
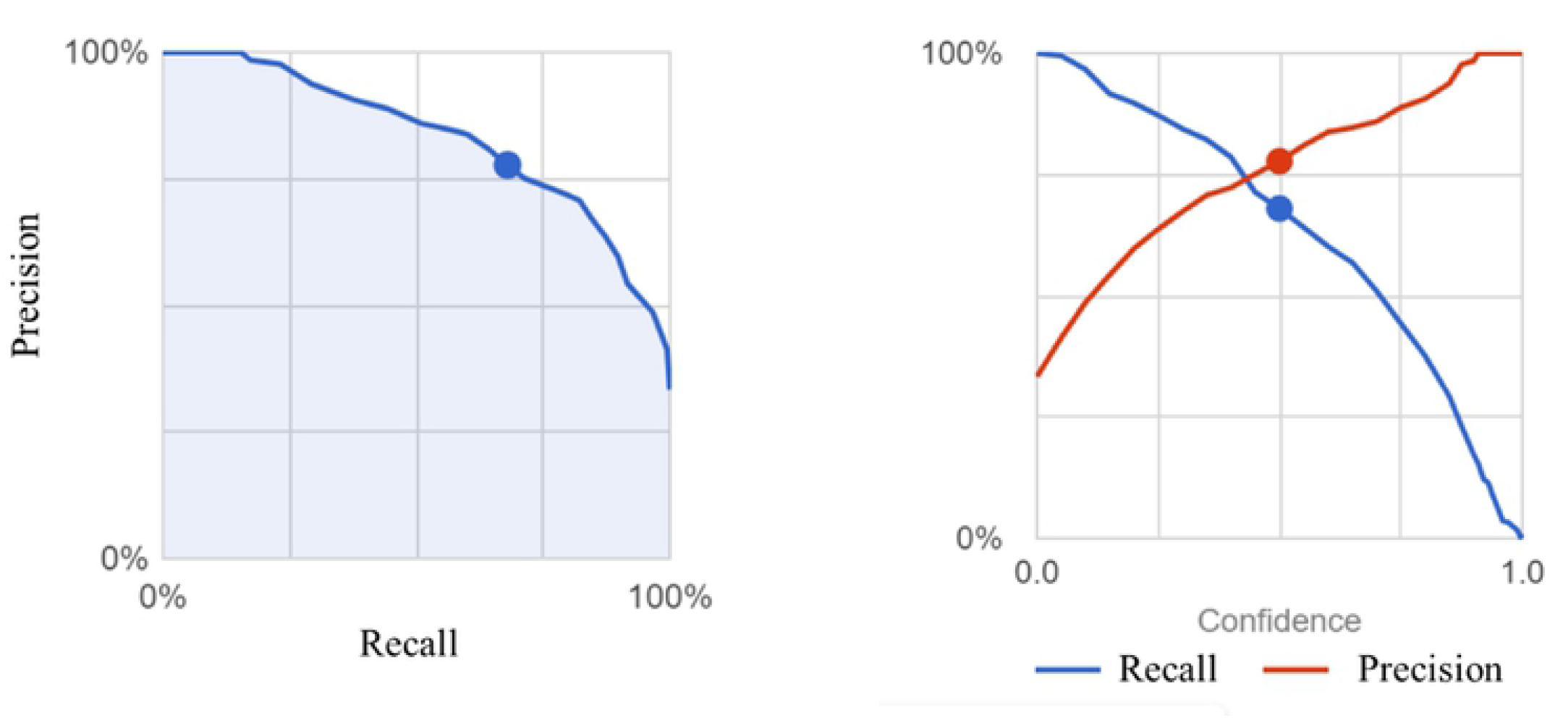
Positive predictive values, recall, and area under the curve (AUC) for a threshold value of 0.5 among healthy, IBS-C, and IBS-D groups.

**Table 2.**
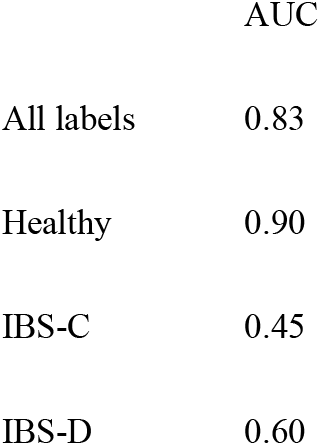
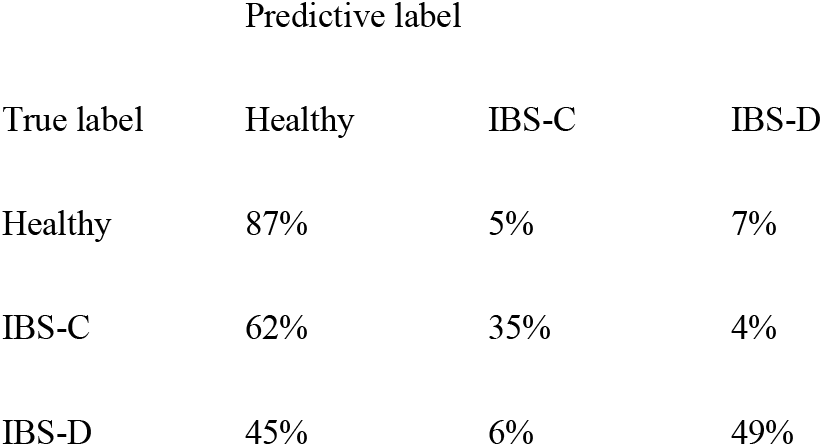
IBS-C, IBS-D and healthy model of the colon; AUC and the contingency tables

**Table 3.**
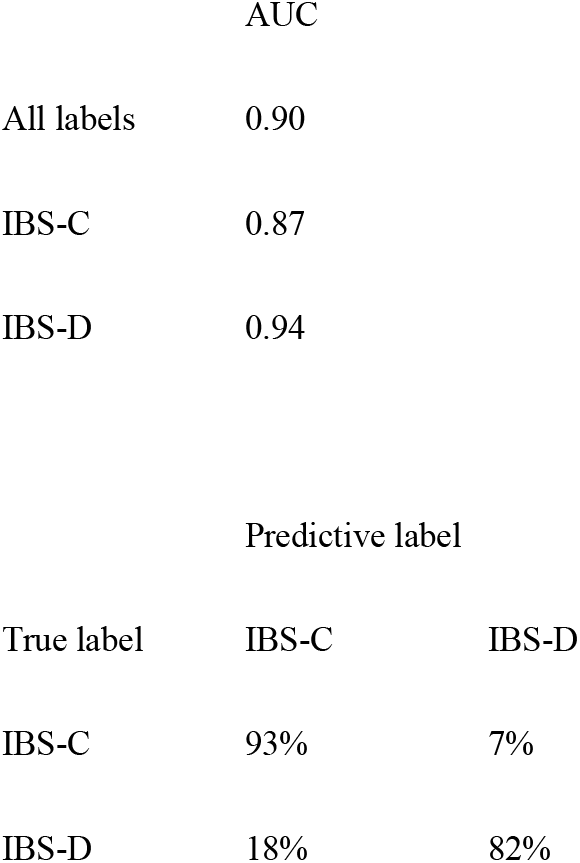
IBS-C, IBS-D model of the colon; AUC and contingency tables AUC

### IBS-C (Group C) vs. IBS-D (Group D)

In comparing Group C and Group D, the average precision (positive predictive value), precision, and recall of the algorithm were 89.75%, 87.5%, and 87.5%, respectively, based on automated training and testing (Figure 5). The precision recall curves and the threshold value were set as described above. The AUC of the model to discriminate among the groups and the confusion matrix are shown in Table 2. The total AUC was 0.90 (0.87 for group C and 0.94 for group D). The confusing rate for group D and group C was 18% and 7%, respectively. Representative images from endoscopy for patients with high IBS-D scores (Figure 6) and high IBS-C scores (Figure 7) are shown.

**Fig. 5.**
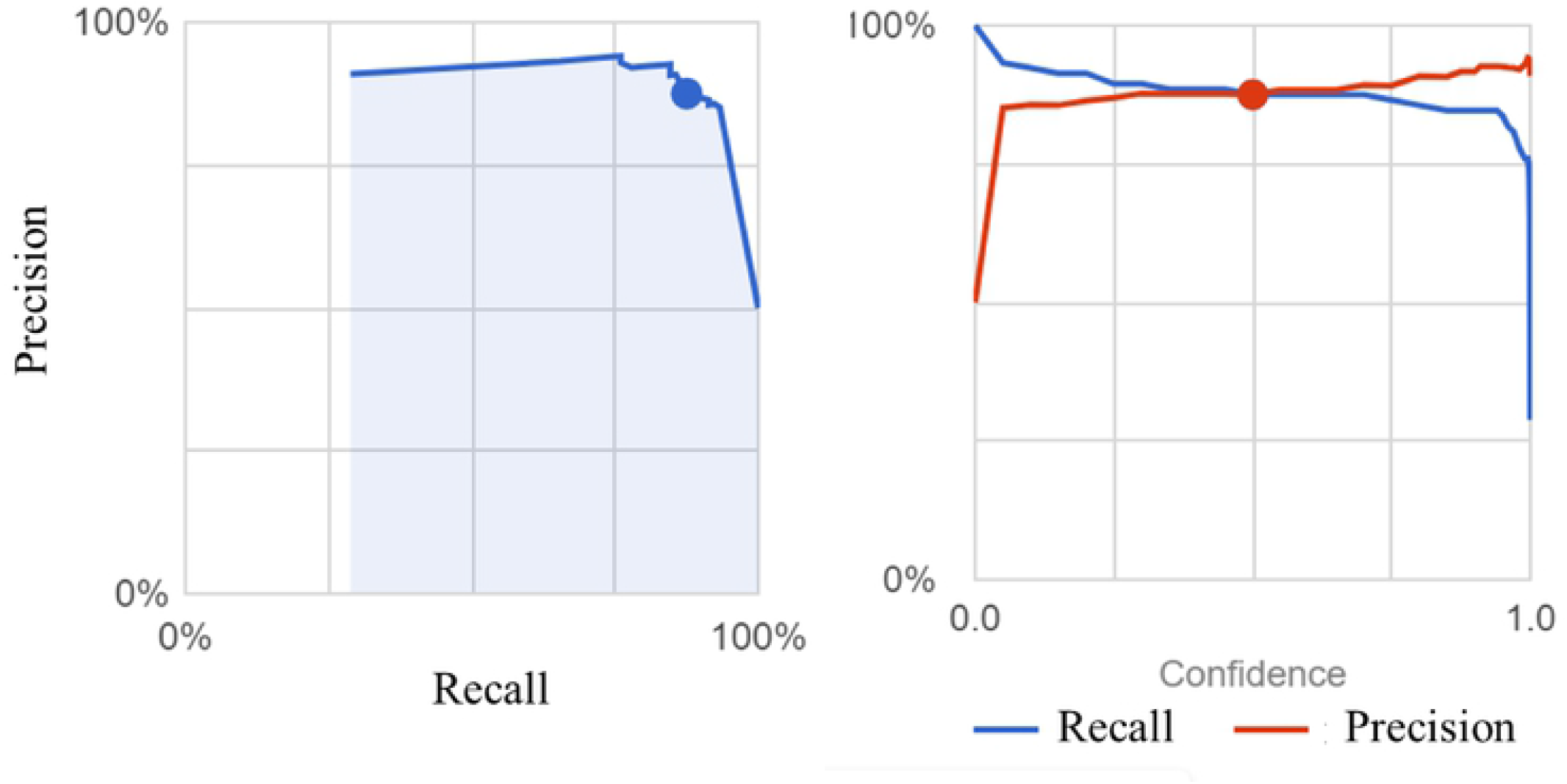
Positive predictive values, recall, and area under the curve (AUC) for a threshold value of 0.5 between IBS-C and IBS-D.

**Fig. 6.**
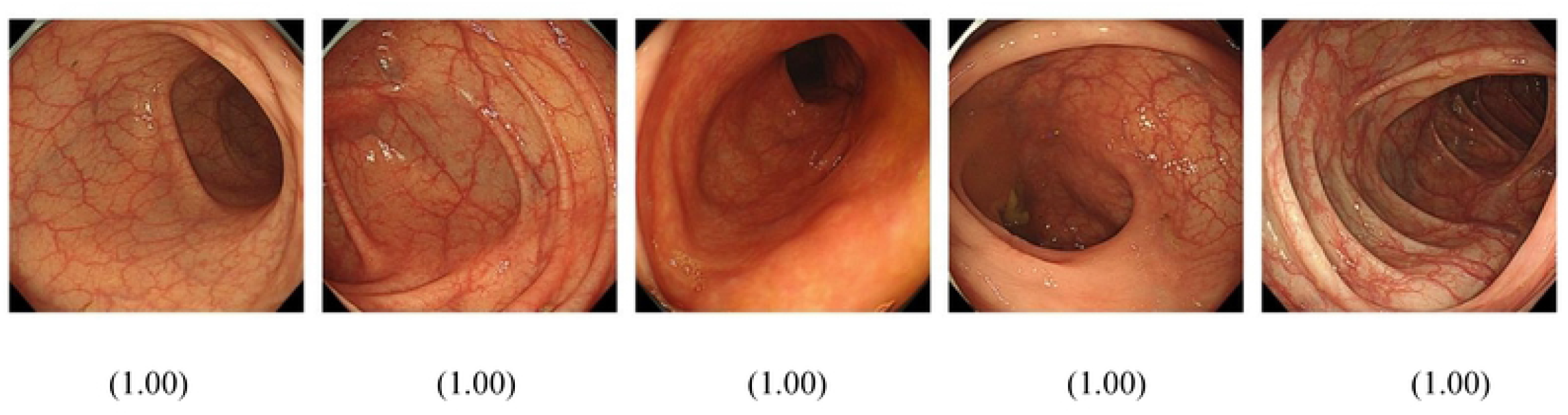
Images that scored high (0 to 1) in the colon image AI model for detecting IBS-D are shown.

**Fig. 7.**
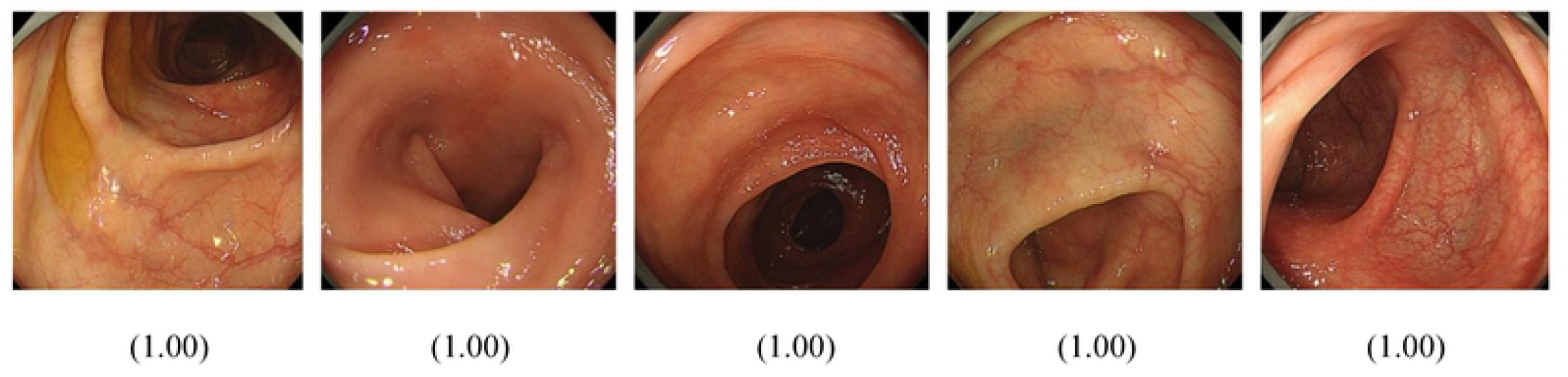
Images that scored high (0 to 1) in the colon image AI model for detecting IBS-C are shown.

## Discussion

Endoscopic images of IBS typically show no abnormalities, but we investigated whether AI could detect microinflammation that cannot be easily detected by human observers. We constructed a code-free AI model to detect IBS with an AUC of 0.95 and a high specificity among healthy subjects. The model was re-created separately for IBS-C and IBS-D, and the AUC was slightly lower at 0.83, suggesting that IBS-C and IBS-D may be distinguishable from each other. When the AI model was reworked to distinguish between the two, the AUC was 0.90, which was greater than the difference between the two groups and healthy subjects. To the best of our knowledge, this is the first AI model that can detect IBS in endoscopic images. Further investigation is needed to determine whether AI can differentially detect histological abnormalities, the presence of biofilm, and/or deformation of the colorectal lumen.

There are several limitations for this model. First, IBS diagnosis was defined not by ROME criteria, but by disease names that were recorded for insurance purposes. However, ROME IV criteria are not always used in clinical practice, and it is thought that the model is rather accurate because it is designed for AI use in clinical practice. Second, patients in the IBS-C (group C) and IBS-D (group D) may have been treated with Linaclotide and Ramosetron, respectively. Third, age, gender, and treatment response were not taken into account when selecting images for the training dataset. Last, the study groups included cases before and after treatment and cases with and without treatment response.

The prime advantage of the use of Google Cloud AutoML Vision is that it requires no coding expertise and can be easily used with datasets to build AI Models. The code-free deep learning approach used in this study has the potential to improve access of clinicians to deep learning[10]. Other research groups have already reported medical image classification and otolaryngology diagnosis that was performed with an automated, coding-free deep learning approach[3, 9, 11]. We have confirmed that AI-based algorithms are also suitable for symptom-based diagnosis. Such algorithms may be able to detect differences in endoscopic images in other functional gastrointestinal disorders, such as functional dyspepsia and nonerosive gastroesophageal reflux disease. The accuracy of the AI model for IBS is expected to vary depending on the light source setting of the endoscope and scope, and whether the same accuracy is achieved at other facilities should be explored. In summary, here we described the development of an AI model that did not require coding experience and that can differentiate IBS, IBS-C, and IBS-D from colonoscopy images. Construction of AI models based on the presence or absence of symptoms could be a new method to diagnose functional gastrointestinal diseases.

## Data Availability

Data cannot be shared for confidentiality reasons. Queries about the data should be directed to the corresponding author.

## Acknowledgements

We thank Ayaka Maeda, Masaya Hiraki, Shun Kuraishi, and Kenji Ogawa, medical engineering technicians at the Medical Device Management Center, University of Toyama Hospital, for their support in collecting and organizing the images. A summary of this study was presented at the 23rd Annual Meeting of the Japanese Society of Neurogastroenterology.

## Ethics

The study protocol was approved by the Ethics Committee of Toyama University (approval No. R2021032). All methods were performed in accordance with the relevant guidelines and regulations, as well as with the Declaration of Helsinki. The study design was accepted by the ethics committee on the condition that a document declaring an opt-out policy by which any potential patients could refuse to be included in the study was uploaded to the Toyama University Hospital website.

## Data Availability Statement

H.M. has ownership of the image data used with Google Cloud AutoML Vision. The data collected during this study cannot be shared.

## Notes

Conflict of Interest Statement The authors have no conflicts of interest to declare.

Funding Sources This work is supported by operating funds from the Mathematical, Data Science and AI Education Program, University of Toyama.

### Competing Interest Statement

The authors have declared no competing interest.

### Funding Statement

The author(s) received no specific funding for this work.

### Author Declarations

The study protocol was approved by the Ethics Committee of the University of Toyama Hospital (Approval No. R2021032). All methods were conducted in accordance with relevant guidelines and regulations and the Declaration of Helsinki. The research plan was presented to the Ethics Committee with the condition that the opt-out policy, which allows patients and their relatives to refuse to participate in the study, is clearly stated on the University of Toyama Hospital website.

